# Relationship between Physical Activity and Self-esteem of High School Students: Cross-sectional Study

**DOI:** 10.1101/2024.01.03.23300489

**Authors:** Zafer Alparslan, Ömür Özer, Eda Nur Arslan, İlknur Irem Aktaş, İbrahım Erdem Susuz, Ayşe Nilüfer Özaydın

## Abstract

**Objective:** This study aimed to investigate the relationship between physical activity and self-esteem of high school students and contribute the literature from a different sociocultural area.

**Methods:** In this descriptive study, whole population of 10^th^ and 11^th^ grade of Capa Science High School (Istanbul/Türkiye) students are invited to study. Voluntarily participating students were asked to answer the questions written on standard survey paper, under the observation of researchers. Survey papers included sociodemographic answers as well as Short Form of International Physical Activity Questionnaire to evaluate physical activity levels and Rosenberg Self-esteem Scale to evaluate self-esteem. IBM SPSS Statistics (Version 11) were used to analyse data. Level of p<0.05 was accepted as statistical significance.

**Results:** Data of 225 persons were succesfully collected (n=278, 80.93%). 16 persons’ data was excluded from physical activity related analyses due to incomplete answers. The 71.60% (n=161) of participants were male, 27.60% (n=62) were female and rest (n=2) did not want to express their genders. Males reported higher self-esteem (p=0.002) and higher physical activity (p=0.031) than females. Self-esteem was associated with regular exercise (p=0.034) status and self-evaluted school success (p=0.001). Self-esteem correlated with participants’ height (p=0.029, r=0.146). Finally, positive correlation between self-esteem and physical activity was found. (p=0.045, r=0.140).

**Conclusion:** Physical activity and self-esteem was correlated with each other in this study. Potential causational relation and underlying mechanisms should be investigated.

## Introduction

Physical activity is one of the key elements that constitute basement of natural and convenient status of one’s life rather than being a temporary enthusiasm. Lack of physical activity does not agree with human body’s physiologic pathways which is being formed through evolutionary period (1).

In accordance with Lieberman et al. stated, “physical activity is just as normal physiological stress as facing of a child’s immune system with pathogen and learning how to cope with it or staying hungry for a while” (2).

There are evidences that show regular exercise delays the onset of 40 different chronic diseases and also increases life expectancy (3). Exercise is likely to enhance quality of life by physical awareness as well as mental and emotional pathways (4).

Although its feasibility and satisfying results on both life quality and disease progress, physical activity trends over years show that it is unlikely to meet the 2025 global physical activity target which was to reduce insufficient physical activity by 10%. Insufficient physical activity shows increase especially for high-income countries citizens (5).

Self-esteem is a reductionist term about how person reflects about himself. It is confidence of one’s own abilites and capabilites (6) and one of the key points of psychological development through childhood and adolescence via its efficacy on favoring psychological stability (7). Also people with higher self-esteem show more life satisfaction (8).

Some of the adverse effects related with lower self-esteem are anxiety, depression and suicidal ideation for school-age children and educational stress can be affecting the self-esteem (9).

In spite of fact that there are researches that show significant association between physical activity status and self-esteem, reviews emphasize the need of more researches from diverse sociocultural areas as mentioned association is likely to be effected by social diversities (10,11). While some studies find positive correlation between physical activity and self-esteem (12,13) some studies do not (14). It is probably due to difference between sociocultural environments of participants. Even the type of high school, science high school, social sciences high school etc., may determine whether there will be relationship between physical activity and self-esteem (14). Also if physical activity has a causative role in enhancement of self-esteem rather than just being correlated with it, social variables are likely to determine this (15).

To consolidate potential relationship between physical activity and self-esteem, unveil whether there is a causative status, figure out how kind of social variables affect this status; literature needs more and more researches from diverse sociocultural environments that yield different social determinants and variables.

In this context, in this study; It is aimed to determine the self-esteem status of 10th and 11th grade high school students and how active they are and to investigate whether there is a relationship between self-esteem and physical activity.

## Methods

### Ethics

i. This study was ethically approved by Marmara University School of Medicine Clinical Studies Ethical Committee. (Protocol code: 09.2023.113/06.01.2023).
ii. The head of high school where the research would be conducted was informed and permission was obtained.
iii. Parents of students under the age of 18 were informed and their consent was obtained.
iv. Each participant was informed separately and their consent was obtained.

### Type of study

This study was a cross-sectional study.

### Participants and study place

The targeted population of study were 10^th^ and 11^th^ grade students of Capa Science High School in Istanbul (N=278). The study sample was not selected and all students were invited to attend to this study.

“Science high schools” in Türkiye are where the most successful students nationwide try to enroll from whole cities with a science exam held by Ministry of National Education. Capa Science High School in Istanbul is one of them. Istanbul is located at the western part of country the most developed and biggest city among the 81 cities in Türkiye.

### Study Process

Flow chart of study is presented in Figure 1.

**Figure 1:**
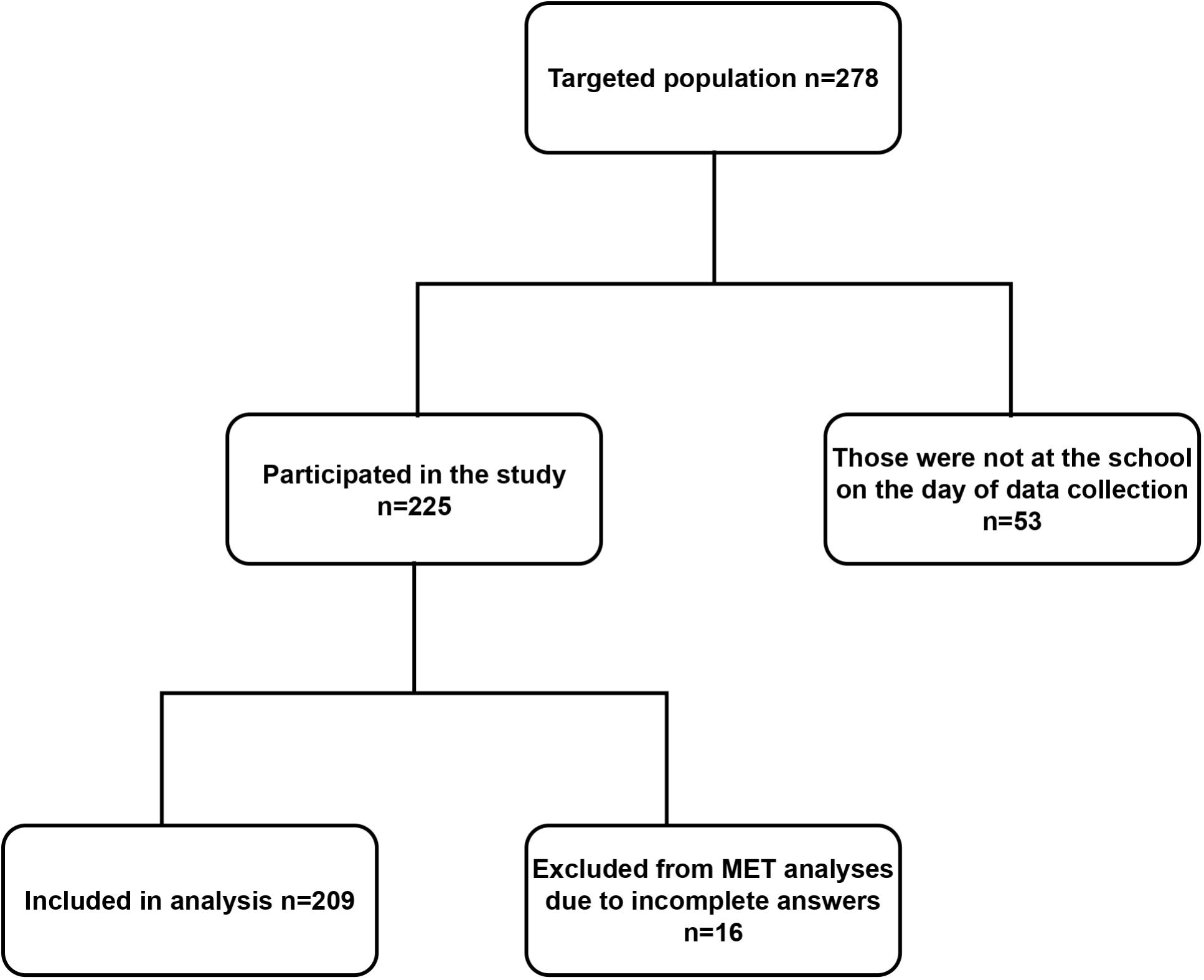
Flow chart of participation and analysis process. There were 278 students at Capa Science High School at grade 10th and 11th in the 2022-2023 school year. Data was collected successfully from 225 students (80.93% of targeted population) who were at school on the day of the study. Answers of 16 participants were excluded from all metabolic equivalent (MET) related analyses according to International Physical Activity Questionnaire (IPAQ) guidelines because of 0 MET points. (Figure 1 legend)

There were 278 students at Capa Science High School at grade 10^th^ and 11^th^ in the 2022-2023 school year. All of them were given both personal and parental consent forms. Data were collected successfully from 225 students (80.93% of targeted population) who were at school on the day of the study. These students had the necessary consent forms already signed by their parents and themselves.

Answers of 16 participants were excluded from all metabolic equivalent (MET) related analyses according to International Physical Activity Questionnaire (IPAQ) guidelines because of 0 MET points.

### Data Collection Tool

The spesific questionnaire for this study was designed by the researcher. This questionnaire has 4 different compartment; first part included the questionnes related sociodemographic characteristics (14 questions). Second part included International Physical Activity Questionnaire Short Form (IPAQ-SF, 7 questions) and third part included Rosenberg Self-Esteem Scale (ROSE, 10 questions). The questionnaire also included some information related the wieght and height of students to be able to calculate and evaluate body mass index (BMI). Based on the statements of the students, it was requested to record the last remembered height and weight values in the last part of the questionnaire.

The questions of sociodemographic characteristics were about birth date, sex, school grade, mother’s education level, father’s education level, regular exercise status, health status based on statement and where they live at.

Participants were asked whether they do regular exercise at school or a club. Participants were asked to evaluate themselves in terms of school success as “not successful, successful, very successful” and school success was assessed according to that.

### Instruments

#### BMI

BMI, formerly called the Quetelet index, is a measure for indicating nutritional status in adults. It is defined as a person’s weight in kilograms divided by the square of the person’s height in metres (kg/m2). The recommended levels are adapted from the global WHO recommendation of 18.5–24.9 as a normal BMI (16).

#### IPAQ-SF

International Physical Activity Questionnaire is a standardized scale which yields both long and short forms to monitor physical activity status developed by researchers from different origins with the aid of World Health Organization and Centers for Disease Control in 2003. Cronbach alpha value is assessed to be between 0.63 and 0.85 (17). Saglam et al. showed reliability and validity of Turkish version of questionnaire in 2010. Turkish version of IPAQ short form (IPAQ-SF) which is reliable and valid is used to determine one’s physical activity status (18). Included physical activities in the IPAQ short form (7 questions) are comprised of both voluntarily exerted physical activities such as carrying weights, running, cycling and also leisure and daily activities such as sitting, transportation-related physical activites. IPAQ-SF yields categorical (ordinal) and continuous scores. Ordinal scores are expressed as inactive, minimally active, HEPA (health enhancing physical activity) active. Continuous scores are calculated as metabolic equivalent (MET) scores. Determination of one’s ordinal score is both affected by both MET and expressed physical activity type (moderate, vigorous etc.) and its frequency. Both ordinal and categorical scores can be used separately or together in order to evaluate one’s physical activity status. Higher MET score indicates higher physical activity, as well. According to IPAQ guidelines, data of participants who had 0 MET score were excluded from all MET related analyses (19).

#### ROSE

Rosenberg Self-esteem Scale, developed by Morris Rosenberg in 1965, is used to evaluate one’s self-esteem. Cronbach alpha value was assessed as 0.86 (20). Validation of Turkish version is provided by Cuhadaroglu in 1986 (21). It is composed of likert-type questions including 4-point response “Strongly disagree, disagree, agree, strongly agree” in the choices. Scale is composed of 10 items and total score can range from 0 to 30. Higher points indicate higher self-esteem status of person. (22).

Data were collected from the students who had parental consent and who agreed to participate in the study, using a survey method under observation on 21^st^ March 2023 in their school.

### Statistical Analyses

Statistical analyses were performed with IBM SPSS Statistics (Version 11). Kolmogorov-Smirnov test was used to evaluate whether data was normally distributed. Mann-Whitney U and Kruskal-Wallis tests were used to check the statistical differences. Spearman’s Rank Correlation is used to evaluate potential correlations. Level of p<0.05 was accepted as statistical significance.

## RESULTS

In this study data was successfully collected from 225 participants (n=278, %80.93).

The 71.60% of participants were male and 27.60% were female. Mean age of male and female participants were 16.50 ± 0.72 and 16.30 ± 0.91 years respectively (p=0.066).

Mean height for male and females were 1.77 ± 0.06 and 1.64 ± 0.05 meter respectively (p=0.001). Mean weight for male and females were 71.19 ± 1.01 and 57.83 ± 1.25 kilograms respectively (p=0.001). Mean BMI for male and females were 22.45 ± 0.26 and 21.37 ± 0.40 respectively (p=0.037).

The sociodemographic characteristics of participants were presented in table 1.

**Table 1:** Sociodemographic characteristics of Participants.

### Rosenberg Self-Esteem Scale (ROSE)

Rosenberg Self-Esteem Scale (ROSE) is a Likert scale where higher points indicate higher self-esteem. ROSE scores were found to be higher for males compared to females (p=0.002). The students’ ROSE scores according to their sociodemographic characteristics were presented in table 2.

**Table 2:** ROSE scores according to sociodemographic characteristics.

Participants were asked whether they do regular exercise at school or a club. The ROSE scores of students who declared that they exercise regularly were found to be higher than those of those who did not (p=0.034), and was shown in graph 1.

**Graph 1:**
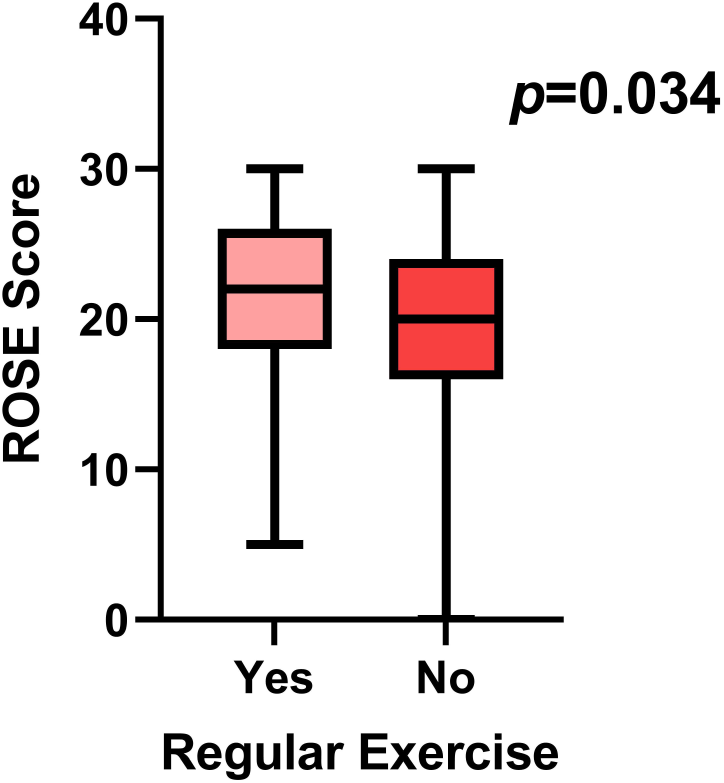
The ROSE scores according to their regular exercise status of students. Participants were asked whether they do regular exercise at school or a club. The ROSE scores of students who declared that they exercise regularly were found to be higher than those of those who did not. Mann-Whitney U test was used to determine significance. (Graph 1 legend)

Moreover, there was positive correlation between ROSE scores and height of participants; it was found that as the student’s height increases, the ROSE score also increases (p=0.029, r=0.146) as was shown in graph 2.

**Graph 2:**
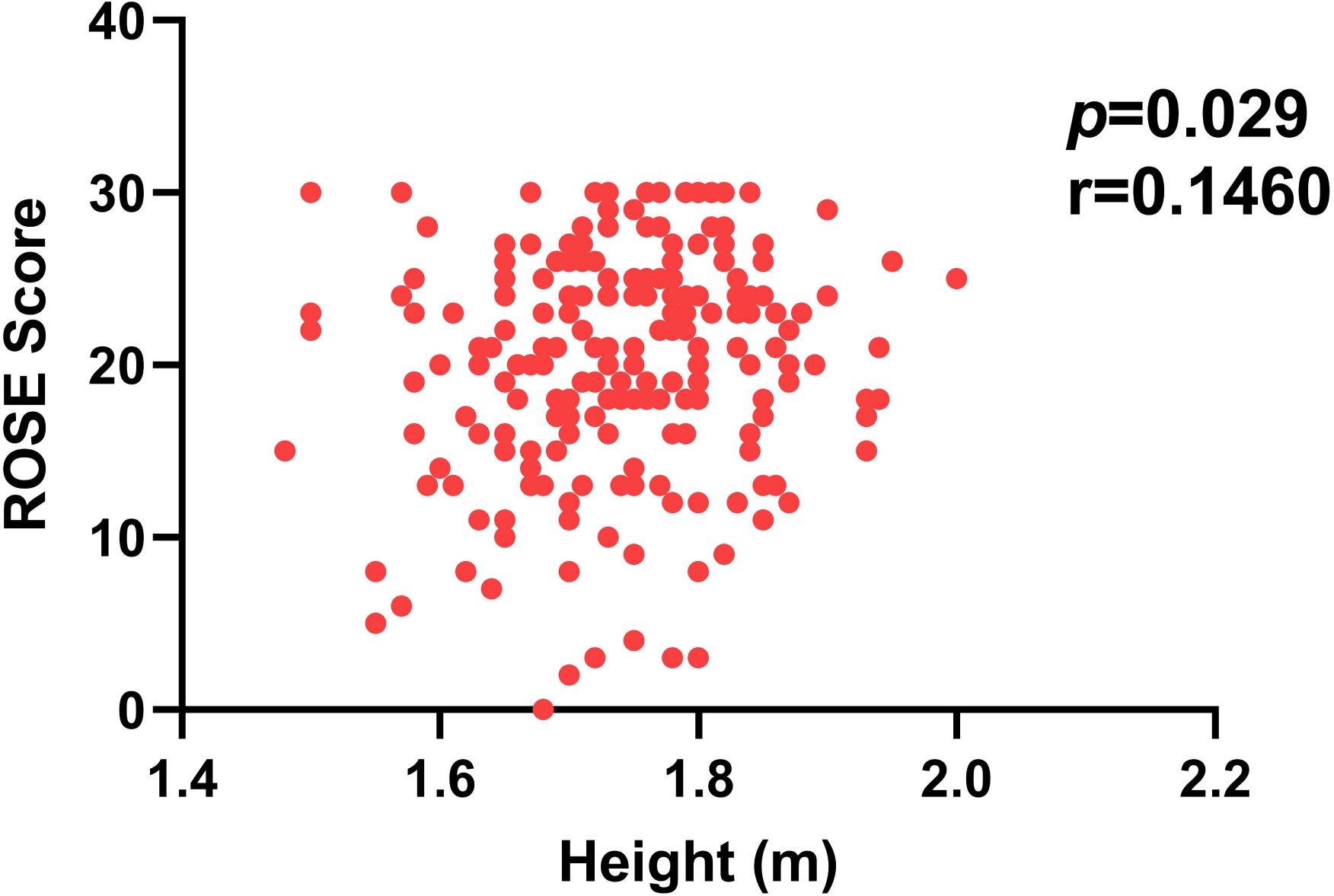
Correlation between ROSE score and height. As height of participants increase, ROSE scores also increase. Spearman’s Rank Correlation was used to determine significance. (Graph 2 legend)

Participants were asked to evaluate themselves in terms of school success as “not successful, successful, very successful”. It has been determined that students with higher school success also have higher ROSE scores than others (p=0.001) as showed in graph 3.

**Graph 3:**
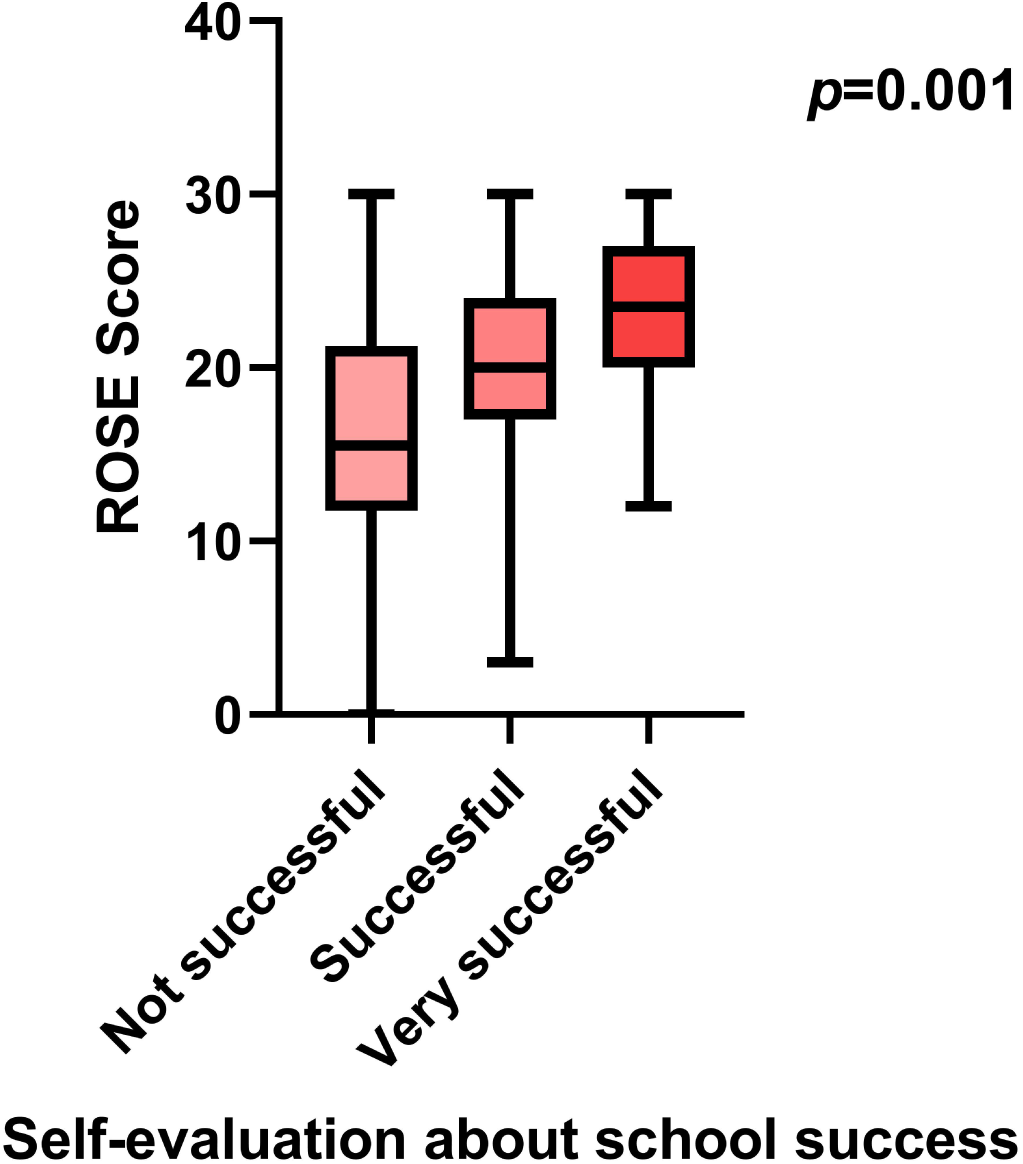
ROSE scores according to school success status of students. Participants were asked to evaluate themselves in terms of school success as “not successful, successful, very successful”. It has been determined that students with higher school success also have higher ROSE scores than others. Kruskal-Wallis test was used to determine significance. (Graph 3 legend)

The 11.20% of participants can be classified as under-weight according to their body mass index (BMI), 70.20% as normal, 16.60% as over-weight, 2% as obese according to data collected by participants’ statement. No differences were found between groups that are separated according to BMI in terms of ROSE scores (p=0.066). Neither differences were found for females (p=0.338) nor men (p=0.124) separately.

The 70.20% of students were normal (according to the WHO classification of their body mass index (BMI) status) but 11.20% of participants under-weight, 16.60% as over-weight, 2% as obese. ROSE scores of students in these four different BMI groups were found to be similar (p=0.066). When this relationship was reanalyzed separately for female and male students, the result was found to be similar (p=0.338, p=0.124).

The 23% of participants were inactive, 61.70% was minimally active and 15.30% were HEPA (health enhancing physical activity) active according to IPAQ-SF.

MET scores as a continuous variable indicating physical activity were derived from IPAQ-SF. It was found that males had higher MET scores when compared to females (p=0.031). Details of the distribution of MET scores based on sociodemographic characteristics were presented in table 3.

**Table 3:** MET scores according to sociodemographic characteristics.

It was noteworthy that MET scores were higher in the group that live in dormitory (p=0.01).

There was positive correlation between ROSE and MET scores (p=0.045, r=0.140) as showed in graph 4.

**Graph 4:** Correlation between ROSE and MET score.

## Discussion

This cross-sectional study aimed to investigate the relationship between self-esteem and physical activity of 10^th^ and 11^th^ grade students of Capa Science High School in 2022-2023 educational year. Study was successfully conducted by visiting school and and questionnaires were answered under the observation of researchers and anonymously. Even though the dramatic earthquakes (7.7 and 7.8 magnitude) which took place on 6^th^ February 2023 in Türkiye affecting at least 11 cities and causing 50.000 people to die, 80.93% of targeted population of this study was successfully reached (23). To evaluate self-esteem and physical activity, scales whose Turkish versions were validated and reliable were used with literature-consistent methods.

There were 62 female participants whereas 161 males. School’s female students are relatively low for every grade because a part of school’s female dormitory was evacuated due to its vulnerability to earthquake, according to school management.

Males reported more self-esteem than females in this study. Generally, studies throughout world report higher self-esteem for men when compared to women. This effect is more prominent for the countries with higher gender equity (24). In line with literature, men had more self-esteem than women in our study. Actually, Türkiye is not a country where gender equality is high, according to Association for Monitoring Gender Equality which is non-governmental organization based in Türkiye. It is ranked as 35^th^ among 36 OECD countries (Organisation for Economic Co-operation and Development) (25). Whereas as mentioned before, our targeted population was very specific that’s why it can differ from the general assesments about Türkiye. Very high exam grades are required to get into the Capa Science High School and also it is located in Istanbul, most developed city of Türkiye. More than half of participants’ mothers and fathers were at least university graduate, those results support our argument about our “specific” population.

ROSE scores were found similar in the groups separated according to BMI. Whereas, a study conducted by Ortega et al. found positive correlation between physical activity and ROSE scores as we have found. They also found negative correlation between BMI and ROSE scores which we have not. Their population age was 10.5±0.5 years (26). Difference between that study and our study could have arised due to ages group difference. What makes us think that is the presence of studies that may indicate BMI’s effect on ROSE scores hinges upon age groups (27,28).

We have found that self-esteem have been positively associated with self evaluation about school success. Zhao et al. found significant association between ROSE scores and both academic engagement and academic self-efficacy. They have measured school-related variables by using scales, respectively Utrecht Work Engagement Scale for Students and Academic Self-Efficacy Scale (29). Even though we have preferred to assess school success without using a scale, our inference that is self-esteem were high for who declared that they were more successful in school is line with mentioned study. There are also papers coherent with our interference indicating that self-esteem and academic achievement/school success is positively associated (30,31).

Participants’ height positively correlated with self-esteem in this study. In the literature association between self-esteem and height is considered to be controversial and there is no consensus. There are some papers showing no significant effect of height on self-esteem (32,33). One of them also alleges that even though actual height of one’s does not affect measured self-esteem; perceptions of their own and people that they communicate about their height may have positive and significant relationship with self-esteem (33). Coherent with that finding, an interesting study using virtual reality by incrementally manipulating participant’s height found that enhancing the height may increase appearance self-esteem (34). Those studies make us think that if height has an effect on self-esteem, the underlying mechanism is dependent on others’ perception. Also, people that preferred limb lengthening surgeries showed higher self-esteem post-operatively (35). There is also a cross-sectional study that finds significant relationship between height and self-esteem in college students (36). Finally, we have found positive correlation between actual height and self-esteem in Turkish high school student adolescents. Importantly, no published articles were found on the internet indicating positive correlation between height and self-esteem for Turkish high school student adolescents by the researchers.

Self-esteem rather than being only positively affected by physical activity can also be playing mediatory role between physical activity and subjective well-being according to Shang et al (37). Physical activity and self-esteem was positively correlated in this study, but mediatory role of self-esteem was not sought.

Dabrowska-Galas et al found positive correlation between ROSE and MET score in a sample including only women (38). In our study, there has been a positive correlation between all participants’ ROSE and MET whereas same is not true for our women participants. Women participants’ ROSE and MET score showed. It could be due to relatively low participant number by women, thus forming a limitation for our study.

As mentioned in the introduction part, even the type of high school may affect how relationship between physical activity and self esteem will be affected. Soytürk et al. found significant association between them in science high school whereas did not find significant association in social sciences high school and sport high school (14). Consistent with those findings, we also have found significant association between physical activity and self esteem in a science high school. Sociocultural determinants play a major role to determine how physical activity affects self esteem or vice-versa.

## CONCLUSION

As a conclusion, we have found that higher physical activity is correlated with higher self-esteem. Males reported more self-esteem than females. Regular exercise was found to be associated with higher self-esteem. Self-esteem was also associated with school success. Participants’ height correlated with self-esteem.

We encourage readers to note those while evaluating the results of this study; this study was conducted in special population. Study was conducted in a science high school where only most successful students all around the country enroll with a special exam. This science high school was located in Istanbul, in western part of counry and most developed city. When those two facts are taken into account together, one can say that our targeted population was very specific special and may show distinct features when compared to rest of country. Also, the information about the height and weights of participants and school success status were collected based on students’ self-reports.

Underlying mechanism and potential causational relationship between physical acitivty and self-esteem waits to be investigated.

## Data Availability

All data produced in the present work are contained in the manuscript

